# The care of older adults with extreme obesity in nursing homes: A collective case study

**DOI:** 10.1101/19013326

**Authors:** Caz Hales, Isaac Amankwaa, Lesley Gray, Helen Rook

## Abstract

**Objective:** To establish the preparedness of nursing homes to deliver high quality, safe and equitable bariatric care for older adults with extreme obesity.

**Design and methods:** A collective case study approach was used. Data collection included observational and interview data from three nursing homes, and a review of 224,200 resident admissions over a 3-year period in New Zealand.

**Participants:** Twenty eight health care workers from three nursing homes in the North Island of New Zealand.

**Results:** Despite a willingness by healthcare staff to care for older adults with extreme obesity, nursing homes were not well equipped to provide safe equitable care for this resident population. Key areas of concern for nursing homes related to limitations in the infrastructure, and financial barriers relating to government funded contracted care services which incorporated equipment procurement and safe staffing ratios.

**Conclusion:** Nursing homes are unprepared to accommodate the existing and increasing number of older adults with extreme obesity who will require bariatric specific care. Government agencies and policy makers will need to consider the financial implications of the increasing need for bariatric level support within aged care, as well as the impact on individual nursing home resources and quality of care provided. Considerable sector and government attention is needed in relation to infrastructure and funding, to allow for the provision of high quality, safe and equitable care for this population group.

## Introduction

The prevalence of obesity increases with age and the numbers of people living with extreme obesity is increasing globally(1,2). The Pacific Islands, United States of America (USA), Saudi Arabia and New Zealand are amongst the countries with the highest rates of obesity worldwide(3), with NZ obesity rates peaking in the 65-74 age group(4); and in adults aged 75 years and over, 24% and 1% are identified with obesity and extreme obesity, respectively. Extreme obesity, as known as bariatric, is typically defined as a body mass index (BMI) of greater or equal to 40 kg/m^2^(5). Within healthcare other clinical definitions of extreme obesity are utilised which assist with care management: weighing ≥150 kgs; having a BMI ≥40 kg/m^2^; or having large physical dimensions which affect mobility and make moving and handling difficult(6).

People with obesity are more likely to develop chronic health conditions earlier in life leading to increasing numbers of individuals with complex care requirements in older age(5). Obesity is known to exacerbate age-related decline in physical function, declining mobility and mobility disability(7). In the older population, extreme obesity has been associated with poorer lower extremity mobility(8,9), with activities such as self-care and moving around physically difficult or impossible(9). Obesity specifically compromises walking, stair climbing and chair rise ability, especially if the BMI exceeds 35kg/m^2^(8,10). Obesity coupled with advancing age increases co-morbidities reduces functional abilities(7), leading to an increase in health service demand and utilisation(11). There is urgent need to understand the preparedness of healthcare services to support this population.

Specific health needs of this population group, regardless of age, include specialised equipment that supports the larger physical dimensions and weight of the person (bed, air mattress, chair, commode, shower chair), specialised moving and handling aids (ceiling and standing hoists, friction reducing devices, grip bars), and increased staff knowledge of specific clinical care needs (hygiene and toileting, nutrition, altered centre of gravity during mobilisation)(6). Additionally, an increase in safety precautions and the need for more human resources has been indicated to prevent adverse health consequences(12).

There is a paucity of research related to older adults with extreme obesity requiring supported care in a residential facility, often referred to as a nursing home, rest home or aged residential care(13,14). These nursing homes provide hospital level care to their resident population. What is currently known relates primarily to the USA population and is intricately linked to their distinctive medical care system(12,14). Whilst more general concerns regarding the principles of care are similar, such as infrastructure and staffing ratios(12,14), understanding the needs and preparedness of nursing homes within other healthcare systems is crucial. To date New Zealand research exploring the extremely obese population has focused on acute hospital management(15–17), and little is known regarding the aged care sector. Whilst the needs of this specific older adult population has increased, service support within the New Zealand aged care sector has largely remained unchanged(18). This paper presents findings of a study examining the requirements of nursing homes to deliver high quality, safe and equitable bariatric specific care for older adults with extreme obesity.

## Methods

The collective case study approach allowed for a holistic exploration and description of the needs of nursing homes to deliver high quality, safe and equitable bariatric specific care in a real-world context(19). A central element of case study methodology is the concept of a ‘bounded system’, the idea being that the boundaries are clear from the outset(19,20). The nursing homes in this research related to one District Health Board (Regional Health Authority) and all received government funded subsidies to support care delivery.

### Study setting and recruitment

The study setting was three nursing homes in the North Island of New Zealand. The three homes were purposively selected because of their unique philosophies and business models. Nursing Home (NH) 1 was a charitable trust, NH 2 a privately listed company and NH 3 was a faith-based organisation. Each home was approached and in turn granted access to the research team. Participant recruitment was purposive(21) and all staff involved in the provision of residents’ care were invited to participate in the study. This included those who delivered regular direct care to the residents and those involved in the operational aspects and resource allocation (nursing home and finance managers).

### Data collection

Four sources of data were used; InterRAI^1^ dataset review, infrastructure spatial measurements, nursing home profile data, and staff interviews. Data collection took place between March and June 2019 with a minimum of one week spent in each home. National and home level InterRAI data was collected from all residents admitted into care between 2015-2018 (224,200 residents). The data provided by InterRAI New Zealand was anonymised. InterRAI analysis was specifically used to identify the prevalence and clinical profile of this resident population.

In preparation for taking spatial measurements, the research team developed and piloted an observational tool to facilitate accurate measurement of space. To ensure accuracy of measurements two members of the research team confirmed each measurement recorded. Existing nursing home equipment was recorded along with specifications including the safe working load. Equipment specifications were validated with equipment providers.

Profile data was gathered at each home and included the number of staff employed, staff to resident ratios, staff turnover, vacancy rates and shift patterns. Nursing home characteristics included occupancy rates, numbers of rest home, hospital and other beds, business models, model of care, the number of premium and standard rooms, cost breakdown of rooms and any additional charges for care. A standardised form for collecting this data was developed and each facility manager completed and validated the information. Twenty-eight nursing home staff were interviewed for the study (Table 1).

**Table 1.** Participant profile.

|  |  |
| --- | --- |
| Participants | N=28 |
| NH 1 | n=10 |
| • Registered Nurse | 4 |
| • Enrolled Nurse | - |
| • Health Care Assistant | 2 |
| • Physiotherapist | 1 |
| • Clinical/Facility Manager | 3 |
| NH 2 | n=9 |
| • Registered Nurse | 3 |
| • Enrolled Nurse | - |
| • Health Care Assistant | 3 |
| • Physiotherapist | 1 |
| • Clinical/Facility Manager | 2 |
| NH 3 | n=9 |
| • Registered Nurse | 2 |
| • Enrolled Nurse | 1 |
| • Health Care Assistant | 4 |
| • Physiotherapist | - |
| • Clinical/Facility Manager | 2 |

Semi-structured interviews were conducted by CH, HR and IA in environments that were negotiated between the interviewer and the participant. Interviews lasted approximately 20-70 minutes. Each interview was audio-recorded and transcribed. Informed written consent was obtained from all participants and data was anonymised with distinguishing participant features removed.

### Analysis

Each of the nursing homes’ profile data, spatial measurements, interview data, and InterRAI data was analysed separately before cross case analysis was conducted to identify the similarities and differences across cases(21). This triangulation of findings enabled a detailed examination of both the phenomena of bariatric care and the context within nursing homes. Best practice guidelines and standards were used to assess the infrastructure and care practices within the homes(6,22–24).

InterRAI data were analysed using SPSS version 24(25). De-identified baseline characteristics of residents were examined by age, sex, ethnic group and underlying clinical condition using descriptive statistics such as proportions and means. Differences in baseline characteristics of the three nursing homes were assessed by one-way analysis of variance (ANOVA) and Chi-square tests for continuous and categorical variables, respectively. Point estimates of individual facility scores for required spaces, equipment and staff were calculated and compared with baseline and demographic data from InterRAI. All statistical tests were conducted at a significance level of p = ≤0.05.

Content analysis was undertaken independently by CH, HR and LG to ensure consistency in the coding of interview data. This approach is regarded as a flexible method for analysing text data as well as being a pragmatic method for the development and extension of knowledge(26). Interviewee responses were aligned to the specific questions asked using the semi-structured interview schedule. Qualitative approaches to ensure the trustworthiness of the study were used with specific attention given to uphold credibility, dependency, confirmability and transferability of findings(27).

## Results

### Overview

The findings from this study highlight that despite a willingness by healthcare staff to care for older adults with extreme obesity, nursing homes were not well equipped or ready to provide safe equitable care for this resident population. Key areas of concern for nursing homes related to limitations in infrastructure and the financial barriers of government funded contracted care services which incorporated equipment procurement and safe staffing ratios. To understand the contextual influences and significance of these care concerns the national prevalence of residents with extreme obesity and the characteristics of the 3 nursing homes are presented.

### 1. National profile of residents with extreme obesity

More residents were overweight, obese and extremely obese than underweight in all three homes (underweight 12.9%, normal weight 46%, overweight/moderate/ severe/extreme obesity 31.6%, missing data 9.5%) and this was comparable to the national trend in BMI ranges across all nursing homes in New Zealand. The national prevalence of residents with overweight, mild/moderate, severe and extreme obesity were 22.1%, 6.7%, 3.7% and 1.1%, respectively. Between 2015-2018 there were 3,811 residents nationally who required some level of bariatric support from their home (Table 2). The heaviest resident cared for within a facility was 264kg (582lbs) and within the 3 homes the heaviest resident ranged from 138-180kg (304-396lbs).

**Table 2.** Description of the anthropometric parameters of residents in the three nursing homes, compared to national figures; InterRAI, 2015-2018.

| Item | Variable | National<br>(N=224,200) | All 3 NHs<br>(N=2,348) | NH 1<br>(n=1,305) | NH 2<br>(n=838) | NH 3<br>(n=205) |
| --- | --- | --- | --- | --- | --- | --- |
| **A** | **Weight (kg)** |  |  |  |  |  |
| 1 | Mean | 67.0 | 62.8 | 62.0 | 63.6 | 62.9 |
| 2 | Min/max | 24-264 | 31-180 | 32-138 | 31-179 | 41-180 |
|  | Missing | 15,855 | 108 | 44 | 55 | 9 |
| **B** | **Height (cm)** |  |  |  |  |  |
| 3 | Mean | 163.5 | 162.7 | 162.5 | 163.6 | 160.7 |
| 4 | Range | 143-213 | 140-194 | 142-198 | 140-194 | 140-182 |
| 5 | Missing | 21,258 | 190 | 130 | 63 | 11 |
| **C** | **Calculated BMI (%)** |  |  |  |  |  |
| 6 | Normal weight | 37.7 | 46.0 | 45.1 | 47.1 | 46.8 |
| 7 | Overweight | 26.6 | 22.1 | 20.2 | 21.8 | 34.6 |
| 8 | Mild/Moderate obesity | 11.3 | 6.7 | 8.2 | 4.5 | 6.3 |
| 9 | Severe obesity | 3.7 | 1.7 | 1.6 | 2.3 | - |
| 10 | Extreme obesity | 1.7 | 1.1 | 0.8 | 1.7 | 1.0 |
| 11 | Missing/not calculated | 9.4 | 9.5 | 10.8 | 8.4 | 5.9 |

### 2. Nursing home characteristics

Operating with different business models, each home had explicit value statements that reflected the nature and purpose of their organisation and included concepts like respect, compassion, holism and excellence. Each nursing home had unique histories for their respective buildings. The charitable and publicly listed homes were purpose-built for the delivery of aged care services. The former facility had benefitted from a recent building extension designed to cater for older adults in need of hospital level care. However, extreme obesity was not considered in the planning stage. Whilst the latter home was outdated and did not meet the specifications that were currently used by the present owners for all newly constructed buildings. The faith-based home was operating from a building that was originally designed for a variety of purposes other than aged care. None of the homes had been purpose-built to accommodate residents with extreme obesity.

The nursing homes sizes ranged from 57 to 153 beds (NH 1, 150 beds; NH 2, 153; NH 3, 57 beds). There was a distinction made between the cost of standard versus premium rooms. In NH 1 all rooms had an ensuite and the size of the room differentiated standard from premium pricing. For NH 2 all rooms were categorised as premium rooms with a cost variance between these premium rooms based on the vista of the room. Nursing Home 3 did not have premium rooms. Each home operated using a volunteer workforce as well as permanent and contracted employees. All homes had rostered and rotating shift patterns with a slight variance between these shifts to allow for differing workplace routines.

### 3. Observations of nursing homes

#### Infrastructure

In all three nursing homes there were infrastructure challenges that hindered the care of older adults with extreme obesity. None of the homes met all the infrastructure standards for bariatric level support. Table 3 provides comparative information on the homes compared to recommended international standards. Specific areas of concern related to door widths which included internal and external doors and emergency exits corridors, and bedrooms and ensuite facilities. There were real issues of safety around the emergency management of older adults with extreme obesity due to the inability to evacuate residents via small doorways:

> *I don’t know whether I should be saying this, but they cannot be taken out from the room-the bed can’t fit through the door [and the resident is immobile]*. (Facility 2, interviewee 8)

**Table 3.** Nursing home infrastructure dimensions compared to recommended standards.

|  | NURSING HOME | RECOMMENDED | STANDARD |
| --- | --- | --- | --- |
| **ACCESSIBILITY: CARPARK** | Carpark space 90°<br>Range 2.5-3.9m width<br>Range 4.2-5.4m length | Carpark space 90°<br>3.5m wide<br>5m length | Worksafe Victoria (2007); International Health Facility Guidelines (2017); Strongwater & Becker (2009); Crook (2009); Standards New Zealand (2001); New Zealand Road Markers Federation (2012). |
| **ENTRANCEWAYS** | Mixed automatic & manual<br>Range 1.3-1.7m width<br>Range 2.0-3.4m height | Automatic doors<br>Minimum width 1.8m<br>Minimum height 2.0m | ACC (2011); Worksafe Victoria (2007); ArjoHuntleigh (2014); Dutta et al. (2018) |
| **EMERGENCY EXITS\*** | Range 1.0-1.4m width | Minimum 1.25m<sup>+</sup> | Collier (2008) |
| **CORRIDORS** | Range 1.2-2.0m width | Wheelchairs 1.2-1.8m width<br>Beds 2.2-2.4m width | ArjoHuntleigh (2014); ACC (2011); Australasian Health Infrastructure Alliance (2009) |
| **FLOORING** | Mixed of carpets, vinyl & slip resistant tiles | Vinyl | VHA (2015); Australasian Health Infrastructure Alliance (2009) |
| **INTERNAL FIXTURES** | Facility 1 & 3 only | Grab bars/handrails especially in corridors | Worksafe Victoria (2007); Strongwater & Becker (2009) |
| **LOUNGES/ COMMUNAL SPACES** | Recliners & standard chairs | 10-20% lounge seating should be bariatric friendly | ArjoHuntleigh (2014); Crook (2009); Gallagher & Steadman (2010) |
| **LIFTS** | Range 1.25-3.6m<sup>2</sup> | *No bariatric specific standards* |  |

\*Excludes main entrances.
<sup>+</sup>Bariatric doors are a recommended minimum 1.5m width.

The sizes of rooms measured fell far below the recommended bariatric dimensions of 25.3m^2^ (28,29). The largest room size measured was 13.9m^2^ (Figure 1). These small rooms created significant logistical and care issues for staff:

> *We had to work out whether it was actually practically possible to get two beds in the room, and then extract one after we’d got her across. So the maintenance guys measured the bed and then went into another room of the same size with two bariatric beds, we’re dealing with wider beds, that don’t fit straight through a doorway. So, they [resident with extreme obesity] have to be on their side, and then lift one sideways within the room. So, they [the staff] did a trial run with two beds. We had to remove all her furniture from the room*. (Facility 2, interviewee 10)

**Figure 1.**
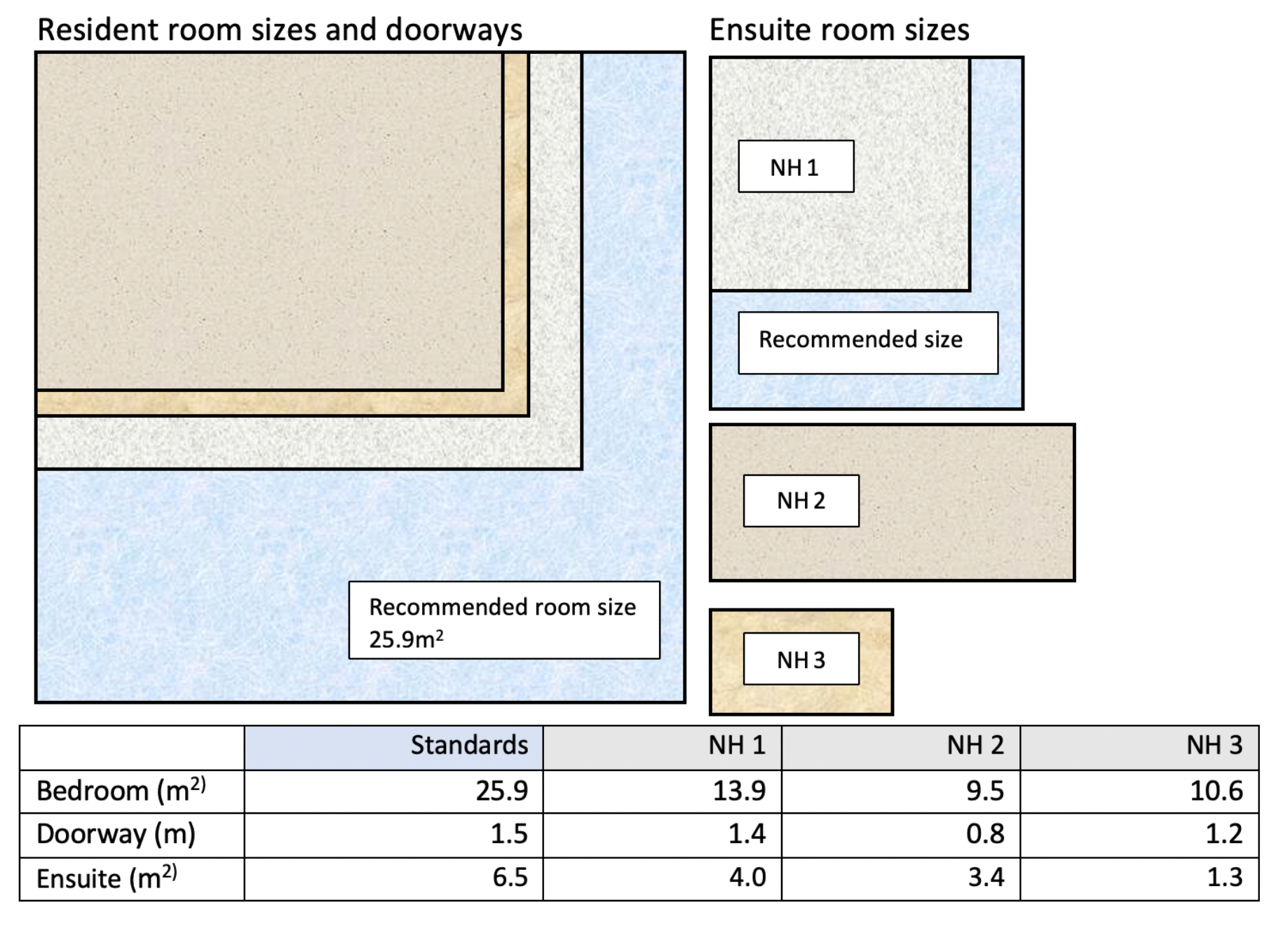
Nursing home room sizes compared to recommended standards.

Ceiling hoists were noted in some rooms however, none of the hoists extended into the ensuite and none were multidirectional and would mean residents with extreme obesity would need to be transferred into a wheelchair prior to moving them into the ensuite. There was poor access to ensuite facilities with door width of 0.8m; the recommended width to allow bariatric access is 1.5m. Overall the ensuite room size ranged from 1.3m^2^ to 4.0m^2^; minimum recommendations are 4.2m^2^. The smallest ensuite was not used for any resident with compromised mobility due to lack of space for healthcare assistants. All homes had toilets positioned close to a corner of the ensuite, restricting movement of residents and caregivers assisting with the care.

#### Equipment

All three homes had the ability to care for fully dependent (non-mobile) residents up to the weight of 120kg without having to procure additional equipment^2^. Beyond 120kg, each home needed to acquire different pieces of equipment and all needed to procure a bariatric bed for any resident weighing over 170kg. For some residents, the physical dimensions of the person could mean a bariatric bed would be required for body weights considerably less than 170kg(15). This could be as low as 130kg. All homes had issues with storage space of the larger pieces of bariatric equipment and there was limited equipment owned by the homes. Often the more costly equipment such as a bariatric bed, was purchased by the resident. Table 4 provides comparative information on the homes’ bariatric equipment space and storage compared to recommended standards.

**Table 4.** Standards for bariatric equipment and storage.

| Type of equipment or space | Recommended consideration | Observed |  |  |
| --- | --- | --- | --- | --- |
|  |  | NH 1 | NH 2 | NH 3 |
| Storage space for bariatric equipment | Facilities may consider creating larger spaces for bariatric equipment or deciding a priori to rent. Whenever possible and safe, equipment frequently used on client can be stored in a client's room, to facilitate access and regular usage(22). | Limited storage space available for bariatric equipment not in daily use. The storage of a bariatric bed would be problematic.<br>The rooms were too small to store equipment; the bariatric wheelchair for a resident was stored at the end of the corridor outside of the resident's room. | Limited storage space for bariatric equipment not in daily use. The storage of a bariatric bed would be problematic.<br>Equipment was stored in the resident's room with other non-essential items removed from the room to create more space. | Limited storage space for bariatric equipment not in daily use. The storage of a bariatric bed would be problematic.<br>Equipment was stored in the resident's room. Although, none had a bariatric bed in use. |
| Friction reduction devices (FRDs) | Examples include sliding sheets, air-assisted devices, beds with turning features and ceiling lifters with repositioning slings.<br>Using FRDs will require a great deal of exertion and expose the caregiver to increased spinal loads. To avoid this, up to 6 caregivers, which is one caregiver per 45kg would be needed(23). | Sliding sheets, ceiling hoists with repositioning slings (SWL 200kg), mobile sling hoist (SWL 227kg) and standing hoist (SWL 200kg) were available.<br>Air assisted devices not available.<br>Staff ratios for moving and handling were reported to range from 2-4 staff.<br>No resident was reported to be over 200kg, in which case more than 4 staff would be required to provide safe care. | Sliding sheets, mobile sling hoists (SWL 227kg) and standing hoist (SWL 200kg) were available.<br>Air assisted devices not available.<br>Ceiling hoists were deliberately not installed, as the provider wanted to maintain a homely environment for residents.<br>Staff ratios for moving and handling were reported to range from 2-4 staff.<br>No resident was reported to be over 200kg, in which case more than 4 staff would be required to provide safe care. | Sliding sheets, ceiling hoists with repositioning slings (SWL 200kg), mobile sling hoist (SWL 227kg) and standing hoist (SWL 200kg) were available.<br>Air assisted devices not available.<br>Staff ratios for moving and handling were reported to range from 2-4 staff.<br>No resident was reported to be over 200kg, in which case more than 4 staff would be required to provide safe care. |
| Bedframes and support surfaces | Bed width may range from 1.0m to 1.4m(49). Preferably, facilities must procure bariatric beds with adjustable width. Additionally, the length of the bed is important, should be able to increase up to 3 meters(29). | Standards beds were 0.9m wide by 2m in length with a SWL 170kg.<br>The standard mattress was 0.82m width by 1.9m length and had a weight capacity of 120kg<br>Bariatric bed and mattress would need to be procured. | Standards beds were 0.9m wide by 2m in length with a SWL 170 kg.<br>The standard mattress 0.82m width by 1.9m length had a weight capacity of 120kg.<br>One bariatric bed and mattress was owned and in use. | Standards beds were 0.9m wide by 2m in length with a SWL 170kg.<br>The standard mattress 0.82m width by 1.9m length had a weight capacity of 120kg.<br>Bariatric bed and mattress would need to be procured. |

#### Finances

Current government funding significantly impacted on the ability of nursing homes to provide equitable care services for older adults with extreme obesity as specified in government-provider contract for services purchased. The services provided by homes covered accommodation, everyday services, core care and support, and allowances for additional care and support. The financial risk for the provider acted as a deterrent for nursing home managers to accept older adults with extreme obesity; safe staffing ratios and equipment procurement were key barriers.

Nursing homes had some flexibility in determining staff to resident ratios based on government service provision agreements that provided recommendations for safe staffing. For hospital level care each resident was allocated 2.4 hours of caregiver and 2 hours of registered time for high acuity residents(30). All staffing costs, however, were expected to be managed under the constraints of the single-bed day payments.

Interviewees in each of the homes discussed staffing ratios as one of the decision-making considerations in caring for older adults with extreme obesity:

> *And then we’d be looking at the staffing, and how we might need to look at the roster [staffing rota] in terms of having the staffing on board for the time that the person would require the cares, and you know at the certain times of the day, to make sure we had enough people on board*. (Facility 3, interviewee 7)

Whilst there was provision for extra financial support to care for bariatric residents, bariatric equipment, a significant financial cost to procure, was considered standard equipment that a provider must supply and therefore not funded under the government-provider contract for services(30). This had a significant impact on individual residents, families, nursing homes and the Regional Health Authority. Facilities often managed with the existing equipment knowing that it was not fit for purpose:

> *Equipment’s an issue, and in this lady’s case we were able to – she already had very large chair, so she already sleeps in her chair all – all night. So, she doesn’t sleep in a bed, so the bed would never ever be wide enough for her, the bed we have at the moment*. (Facility 1, interviewee 6)

Residents and family were often expected to make additional financial contributions to care needs because of a resident’s larger size and the financial implications for continued care:

> *Sometimes the families, when they’re bringing in someone that is obese, sometimes they come with the equipment*. (Facility 1, interviewee 7)

This additional resident charge is contrary to the New Zealand healthcare system which primarily operates a free health service for users, with some long-term care services, such as residential care, means tested with a maximum daily contribution.

## Discussion

The key findings of this research identified that whilst healthcare staff are willing to care for older adults with extreme obesity, the nursing homes were not well equipped or ready to provide safe equitable care for this resident population. Key areas of concern for nursing homes related to limitations in the infrastructure of current facilities, and financial barriers that specifically related to equipment procurement and safe staffing ratios. Such concerns have been well documented in the USA in relation to increasing prevalence of obesity in aged care facilities(31–34). A systematic review of 28 studies examining the impact of obesity on nursing home care in the USA identified that obesity posed major issues for the provision of nursing home care, impacts the wider health care system, and provides challenges for nurses to deliver high quality care (35).

Our findings indicate the need to prepare nursing homes to receive and provide safe and equitable care for the increasing obese population. Challenges and barriers in providing safe equitable care were partly because of outdated infrastructure, developed prior to the need for bariatric specific care services. Nursing homes in this study were not purpose built and have been remodelled and retrofitted. Although the adaptations meet the care needs of most residents, none had areas specifically designed for older adults with extreme obesity. This is not unexpected given that infrastructure specification standards are relatively new for the care of bariatric people in New Zealand. Similar issues with inadequate infrastructure have been identified in other countries who are having to consider the costs and issues of retrofitting healthcare facilities to accommodate the specific needs of the bariatric population(36). Renovating existing structures to make them ‘bariatric friendly’ appears to be the most pragmatic step for nursing home managers to take(36). This may involve remodelling and retrofitting existing buildings to provide extra space. Given the space constraints and high resources required to set up newer facilities, it is pertinent for healthcare planners to renovate the existing homes to make them suitable for this population.

Existing infrastructure poses serious safety concerns regarding the evacuation of residents with extreme obesity during emergency situations. These concerns were primarily related to the width of designated emergency exit doors in the resident care areas. Concerns about the limitations of healthcare infrastructure in the emergency evacuation of those with extreme obesity have been highlighted in accounts of emergency management processes following disasters(37). Issues related to how the physical size of the person prevented the use of evacuation equipment fitting through narrow stairwells(38,39), and safety concerns for the person and the rescue staff should the person fall(38). In situations where power failures may occur, facilities must consider the needs of people reliant on electric devices and have adequate arrangements for backup power(40–42). Additionally, it has been suggested that consideration needs to be given to factoring in the predicted longer evacuation time of larger and older adults(43). Based on our findings, it is imperative that homes develop emergency management plans and procedures that consider the weight and size of residents.

The funding model in New Zealand was developed in the 1990s and based on the resident clinical profile of that time. The average pricing approach of the 1990s makes it difficult for providers to manage the higher care costs associated with bariatric residents. Whilst there is provision for extra government financial support to care for bariatric residents, bariatric equipment, a significant financial cost to procure, was not part of the eligibility criteria for additional funding. Therefore, the lack of bariatric equipment owned by the homes was identified as a barrier to admission. This issue of equipment is not unique to New Zealand nursing homes, a study by Bradway et al.(44) indicated that 81% of nursing homes lacked the necessary bariatric equipment; the existence of ‘equipment concerns’ was associated with reporting extreme obesity as a barrier to admission from a hospital setting. These barriers to accessing nursing homes are common(31,44) and raise significant concerns regarding the equity of service provision for older adults with extreme obesity. In our study, older adults with extreme obesity were either declined admission or residents and their families carried the financial burden of purchasing essential care equipment.

Staffing ratios for residents who require hospital level care was a financial barrier to accepting older adults with extreme obesity. The funding for 2 hours of registered nurse and 2.4 hours of health care assistant support per resident with high acuity needs, per day was not sufficient to safely care for this resident population. When assessing mobilisation needs practice standards recommend one caregiver per 45kg of patient weight when using safe moving and handling equipment (friction reduction devices i.e. sliding sheets)(23). When applying these standards to future bariatric residents it is expected that staffing ratios or staff time allocation needs to be considerably higher. As older people’s BMI increases caregivers need more time than when they perform the same task with non-obese residents(31,45–47). Kosar et al.(47) examined whether different levels of obesity was associated with higher staffing needs for completing activities of daily living (ADL) amongst care home residents. Higher obesity levels was significantly associated with assistance from 2 or more staff for all ADLs except eating. With increasing prevalence of obesity in nursing homes, funding models for staff resources that are not adjusted for extreme obesity or do not account for additional ADL assistance need to be amended.

Findings from this study offer the global aged care sector insights into challenges facing the aged care sector in New Zealand. The study draws upon a large quantitative national dataset along with data from three nursing homes with different business models and philosophies. A limitation of this study is that the Central Region’s Technical Advisory Services Limited in New Zealand gives no warranty that the information supplied in the InterRAI system is free from error. However, studies from other countries utilising InterRAI datasets report consistent and stable trends in internal consistency(48). A further limitation is that New Zealand has its own funding model for aged care provision, and this might not directly translate to other countries in the same way. However, international research supports the findings within this report.

## Conclusion

This study is the first of its kind in New Zealand and highlights that nursing homes are not prepared to accommodate the existing and increasing number of older adults who will require bariatric level care. Based on this study and other international research, it is crucial that government agencies and policy makers consider the financial implications of the increasing need for bariatric level support within aged care, as well as the impact on individual nursing home resources and quality of care provided. Significant investment is needed to address the safety, equity and care concerns of older adults with extreme obesity at government and organisational levels with considerable attention to infrastructure and funding. Further research is needed to understand projected funding requirements of the aged care sector to meet infrastructure and care demands.

## Data Availability

No data is publicly available

## Acknowledgements

We would like to thank the participants in the study and acknowledge the advisory group who guided specific aspects of the research: Alyson Kana, Senior Policy Analyst, New Zealand Aged Care Association; Pakize Sari, General Manager of Te Hopai Home and Hospital, Wellington; Perry Robertson, NZ Operations Manager and Todd Bishop, Company Director, Essential Helpcare, Healthcare equipment provider; Dr Lisa Te Morenga, Senior Lecturer in Māori Health, School of Health, Victoria University of Wellington.

## Funding

This study was fully funded by a University Research Fund, Victoria University of Wellington, New Zealand.

## Conflicts of Interest

No conflicts of interest

## Disclaimer

The views expressed are those of the authors and not the New Zealand Aged Care Sector

## Ethical Approval

Granted by Victoria University of Wellington Ethics Committee (Approval Number 27169). Additionally, approval was sought from the Central Region’s Technical Advisory Services (TAS) to access the InterRAI dataset.

## Footnotes

1 A nationwide mandatory clinical assessment tool used in all nursing homes for all residents in New Zealand.

2 The room size would still be inadequate to safely manoeuvre the mobilisation equipment and accommodate the necessary extra staff.

## Notes

### Competing Interest Statement

The authors have declared no competing interest.

